# Immunogenicity and safety of an inactivated SARS-CoV-2 vaccine (BBV152) in children from 2 to 18 years of age: an open-label, age-de-escalation phase 2/3 study

**DOI:** 10.1101/2021.12.28.21268468

**Authors:** Krishna Mohan Vadrevu, Siddharth Reddy, Harsh Jogdand, Brunda Ganneru, Nizam Mirza, V.N. Tripathy, Chandramani Singh, Vasant Khalatkar, Siddaiah Prasanth, Sanjay Rai, Raches Ella, William Blackwelder, Sai Prasad, Krishna Ella

## Abstract

**Background:** We assessed the safety, reactogenicity, and immunogenicity of BBV152 in an open-label age de-escalation study in three age cohorts of children from 18 years of age down to 2 years of age.

**Methods:** This was a phase 2/3 open-label, multi-centre study done across six hospitals in India. All children received two 0.5mL doses of BBV152 (Covaxin^®^, Bharat Biotech International Ltd., Hyderabad, India), which is the same formulation indicated in adults. Participants were monitored for adverse events, and post-vaccination blood draws were collected to assess neutralising antibodies. A total of 526 children were enrolled into Group 1 (ages 12 through 18 years, n=176), Group 2 (ages 6 through 12 years, n=175), Group 3 (ages 2 through 6 years, n=175).

**Findings:** There were no serious adverse events, deaths, or withdrawals due to an adverse event during the study. Vaccination with BBV152 was generally well tolerated, with no substantial difference in reactogenicity profiles between the different age groups. Similar immune responses were measured as microneutralisation (MNT) antibody titers in all three age groups. Vaccine-induced MNT responses in all groups were comparable to BEI reference sera run in the same assay. Seroconversion (measured by Plaque Reduction Neutralization Test (PRNT)) achieved high levels (95-98%) in all three groups four weeks after the second vaccination. The PRNT GMT ratio was 1·76 (95%CI: 1.32 – 2.33) (GMT all children subgroup / GMT in adults) had a lower limit ≥ 1, indicating superior antibodies in children when compared to adults. Vaccine responses were skewed towards a Th1 response with IgG1/IgG4 ratios above 1.

**Interpretation:** BBV152 is well tolerated and immunogenic in children from 18 years down to 2 years of age. Immunogenicity analysis (by PRNT) shows superior antibody responses were observed in children compared to adults, suggesting that BBV152 will also be efficacious in this age group.

## INTRODUCTION

There is an ongoing unmet medical need for effective vaccines against the SARS-CoV-2 virus to combat the global COVID-19 pandemic. This is particularly relevant in low- and middle-income countries which can neither afford nor provide the necessary infrastructure to store and distribute the most widely used vaccines, which require low temperatures [1]. To meet this need, Bharat Biotech has developed BBV152 (COVAXIN^®^), a whole virion adjuvanted inactivated SARS-CoV-2 vaccine that is stored at standard refrigerator temperatures (2–8°C) [2]. In a trial involving over 25,000 adults, we have recently demonstrated that two doses of BBV152 have 77.8% efficacy against RT-PCR confirmed COVID-19 of any severity [3]. A smaller study in 3732 Indian healthcare workers in conditions of high infectious pressure during the second wave of infections, probably of the Delta variant, found effectiveness of 47–57% depending on the interval since vaccination [4].

Paediatric COVID-19 infections have not been associated with the high rates of morbidity and mortality observed in older adults and paediatric deaths due to COVID-19 are rare, typically occurring in children with underlying chronic medical conditions [5,6]. Children have also been shown to have similar viral loads as adults during infections that tend to be asymptomatic and may be a source of the ongoing infections in more susceptible populations [7]. However, the increasing occurrence of Variants of Concern [8], which have almost entirely replaced the original prototype virus, may lead to higher rates of severe disease in children. In addition, the ongoing pandemic and failure to decrease rates of infections have been associated with an increasing proportion of infections being in children [9]. This has led several countries to consider extending their current adult vaccination programmes to include children [10,11], making the development of effective vaccines or the establishment of current vaccines as being suitable for use in children a medical priority, to both protect this population and to increase herd immunity. To meet this need, we assessed the safety, reactogenicity and immunogenicity of BBV152 in an open-label age de-escalation study in three age cohorts of children from 18 years of age down to 2 years of age.

## METHODS

### Study design and participants

This was a phase 2/3 open-label, multi-centre study done across six hospitals in India. The protocol was approved by the Institutional Ethics Committees at each study centre and the study was done in accordance with the revised Declaration of Helsinki (64th WMA General Assembly, Fortaleza, Brazil, October 2013). This report presents data obtained up to four weeks after the second of two vaccinations, but the study is ongoing to collect data up to six months after the last vaccination, which will be reported separately. Eligible participants were healthy children of either gender from 2 to 18 years of age whose parents or legal guardians supplied written informed and audio-video consent; verbal assent was also obtained from children between 7 and 12 years of age, and written consent from children >12 to 18 years of age. A Data Safety Monitoring Board (DSMB) comprising independent medical and vaccine experts was convened to assess the safety of the vaccinations throughout the study and to approve the age de-escalation.

Inclusion criteria included general good health in the investigator’s opinion, ability to fulfill the study criteria, to remain in the study area for its duration, and not to participate in any other clinical trial. Main exclusion criteria were any history of prior COVID-19 vaccination, a SARS-CoV-2 infection confirmed by RT-PCR or ELISA at screening, a temperature (>38·0ºC) or symptoms of an acute illness within 3 days of a vaccination, known sensitivity to any vaccine component, receipt of any other vaccine within 4 weeks of the study or any know immunosuppressive condition or treatment likely to interfere with the immune response.

The vaccine antigen is a β-propiolactone-inactivated whole virion of vaccine strain NIV-2020-770 sourced from the Indian Council of Medical Research National Institute of Virology (ICMR-NIV; Pune, India). Each 0.5mL dose of BBV152 (Covaxin^®^, Bharat Biotech International Ltd., Hyderabad, India) contains 6 μg antigen with a toll-like receptor 7/8 agonist molecule (imidazoquinoline; IMDG) adsorbed to alum (Algel-IMDG). Vaccine was supplied in prefilled syringes, stored at 2–8°C, and was administered by intramuscular injection in the upper deltoid.

### Procedures

Age de-escalation was done by first recruiting Group 1 with the 18 to <12 years age cohort. The first 25 participants received their first vaccination, and 7 days of safety and reactogenicity data was then assessed by the DSMB before it gave approval to recruit and vaccinate the remaining 150 participants in that age group. When the vaccination of Group 1 was complete, this process was repeated with Group 2 aged > 6 to 12 years, the first 25 being vaccinated and assessed after 7 days, before the remaining 150 were vaccinated. Finally, Group 3, aged ≥ 2 to 6 years was recruited, the first 25 were vaccinated and assessed for 7 days before the remaining 150 were vaccinated.

All children were screened for COVID-19 by RT-PCR and ELISA 1–4 days before their first vaccination on Day 0, when eligible participants had a general physical examination and assessment for potential COVID-19 infection before the first blood draw. The vaccine was then administered, and the participant was monitored for two hours for immediate reactions. Parents/guardians were trained on how to complete study diaries for 7 days in which were solicited local reactions and systemic adverse events. The solicited local reactions (pain, redness, swelling, stiffness, and tenderness) and systemic AEs (body pains, fatigue, headache, loss of appetite, nausea, vomiting, weakness, and fever) were graded for severity as mild (aware but easily tolerated), moderate (discomfort sufficient to interfere with normal activity) or severe (incapacitating and unable to perform normal activity). Active surveillance of reactogenicity and possible COVID-19 infection was also done during these 7 days through telephone contact with the parents/guardians who were instructed to record any unsolicited adverse events which occurred before the next study visit, and to report immediately to the investigator any serious adverse event (SAE) or any adverse event of special interest (AESI). AESIs included anaphylaxis, vaccine-associated enhanced respiratory disease (VAERD), generalised convulsions, acute respiratory distress syndrome (ARDS), and pneumonitis.

This process was repeated on Day 28; a blood sample was drawn before participants received a second vaccination and parents/guardians recorded solicited local and systemic adverse events for 7 days with daily telephone contact. At a third visit on Day 56 a third blood sample was drawn to assess immune responses.

### Immunogenicity

Sera were prepared immediately from all blood draws and kept at -20ºC for shipping to the Bharat Biotech International laboratory (Hyderabad, India) for measurement of immune responses using microneutralisation (MNT) and plaque reduction neutralisation (PRNT) tests, with titres expressed as reciprocal of the dilution which achieved 50% neutralisation (MNT_50_ or PRNT_50_). Due to the lack of an established correlate of protection against SARS-CoV-2 infection, vaccine-induced responses were compared with an internationally recognised reference serum (Biodefense and Emerging Infections Research Resources Repository, NIAID, NIH). We also measured the titres of IgG antibodies against the S-protein, receptor-binding domain (RBD), and nucleocapsid protein (N-protein) of SARS-CoV-2 by ELISA expressed as arbitrary ELISA units/mL. A post hoc analysis of randomly selected serum samples (n = 24) was done to measure IgG1:IgG4 ratios as an indicator of the balance of the Th1:Th2 responses [12,13].

### Statistical analysis

The primary safety and reactogenicity objectives were to assess the occurrence of solicited adverse events (AE) within 7 days of a vaccination, unsolicited within 28 days of a vaccination, and serious AEs (SAE) and AEs of special interest (AESI) throughout the study duration. The primary immunogenicity objective was to assess the geometric mean titres (GMTs) and seroconversion rates of neutralising antibodies. Safety and reactogenicity comparisons were made descriptively between groups as group proportions reporting any and each specific adverse event.

Immunogenicity analyses were to be done on all those enrolled and treated. Per-Protocol non-inferiority (NI) analyses were based on those who received both doses of vaccine. The sample size was intended to allow separate comparisons of geometric mean neutralisation titres at Day 56 between each of the three age groups, and against titres observed in adults in a phase 2 study [14].

The non-inferiority criterion for adequacy of the immune response in the paediatric population used an NI margin of 0.5. Thus, a paediatric age subgroup was considered non-inferior to the adult group if the 95% confidence interval (CI) for GMT ratio (GMT in a paediatric subgroup / GMT in adults) had a lower limit ≥ 0.5. Based on the adult phase 2 trial data, we assumed that the standard deviation of log_10_ (titre) is 0.5. To be conservative, we assumed the true GMT ratio is 0.8 for each paediatric subgroup – i.e., that the true underlying GMT is lower in the paediatric subgroup. We assumed sample sizes of 150 individuals providing data for each paediatric subgroup and 177 for the adult group, which is the number reported from the phase 2 trial with titre data at Day 56. Using a t-test for non-inferiority on the difference in means of log_10_ (titre) with assumed true difference of log_10_ (0.8) = -0.096910 and NI margin of log_10_(0.5) = -0.30103, the power to show non-inferiority for a single paediatric age subgroup is approximately 0.95615 (PASS 2020, NCSS, Kaysville, Utah, USA). Then the probability that the NI criterion would be met for all three paediatric age subgroups is approximately (0.95615)^3^, or 87%. To allow for loss of data due to withdrawals, loss to follow-up, etc., we planned to enroll 175 children into each of the three paediatric age subgroups.

The GMT was calculated for neutralisation titres and for ELISA IgG titres for each age group separately. A two-sided 95% confidence interval (CI) for the postvaccination GMT was calculated from a 95% CI for the mean of log-transformed titre, using a normal approximation for the distribution of log (titre). For each pair of age groups, the ratio of GMTs and the corresponding 95% CI are presented. The 95% CI for the GMT ratio was calculated from a 95% CI for the difference in means of log_10_ (titre). Seroconversion rates (SCR) were calculated as group proportions achieving a four-fold increase in titre from baseline (Day 0) to Day 28 or Day 56; two-sided 95% Cis were calculated for SCRs. For differences between SCRs, exact 95% CIs were calculated, and the SCRs were compared using two-sided Fisher exact tests, with the p-value obtained by doubling the smaller of the one-sided p-values.

### Role of the funding source

The sponsor of the study had no role in data collection, data analysis, data interpretation, or writing the report. A contract research organisation (CRO) was responsible for data analysis and generating the report. The unblinded CRO and several authors from Bharat Biotech had full access to the data in the study. Authors from Bharat Biotech had final responsibility for the decision to submit for publication.

## RESULTS

From 26 May to 10 July 2021 we screened 976 potential participants of whom 342 were excluded for seropositivity for COVID-19 according to the RT-PCR or ELISA testing (**Figure 1**). Of the 634 eligible children 526 were enrolled and vaccinated, 176 in Group 1 and 175 in Groups 2 and 3. Only one child did not receive their second vaccination – a participant in Group 1 who was diagnosed with COVID-19 on Day 15. As shown in **Table 1** there was an imbalance of males over females in each age group, this difference is the largest in the youngest group.

**Table 1:** Baseline characteristics

|  | **Group 1 (n = 176)**<br>> 12 to 18 years | **Group 2 (n = 175)**<br>> 6 to 12 years | **Group 3 (n = 175)**<br>≥ 2 to 6 years |
| --- | --- | --- | --- |
| **Age, years** | 14.52 (1.59) | 9.01 (1.67) | 3.72 (1.10) |
| **Sex** |  |  |  |
| Female | 84 (47.7) | 72 (41.1) | 68 (38.9) |
| Male | 92 (52.3) | 103 (58.9) | 107 (61.1) |
| **Weight, kg** | 55.5 (13.7) | 34.3 (9.8) | 16.4 (3.7) |
| **BMI, kg/m<sup>2</sup>** | 21.1 (3.9) | 17.6 (2.8) | 15.4 (2.6) |

Data are Mean (SD) or n (%)

**Figure 1.**
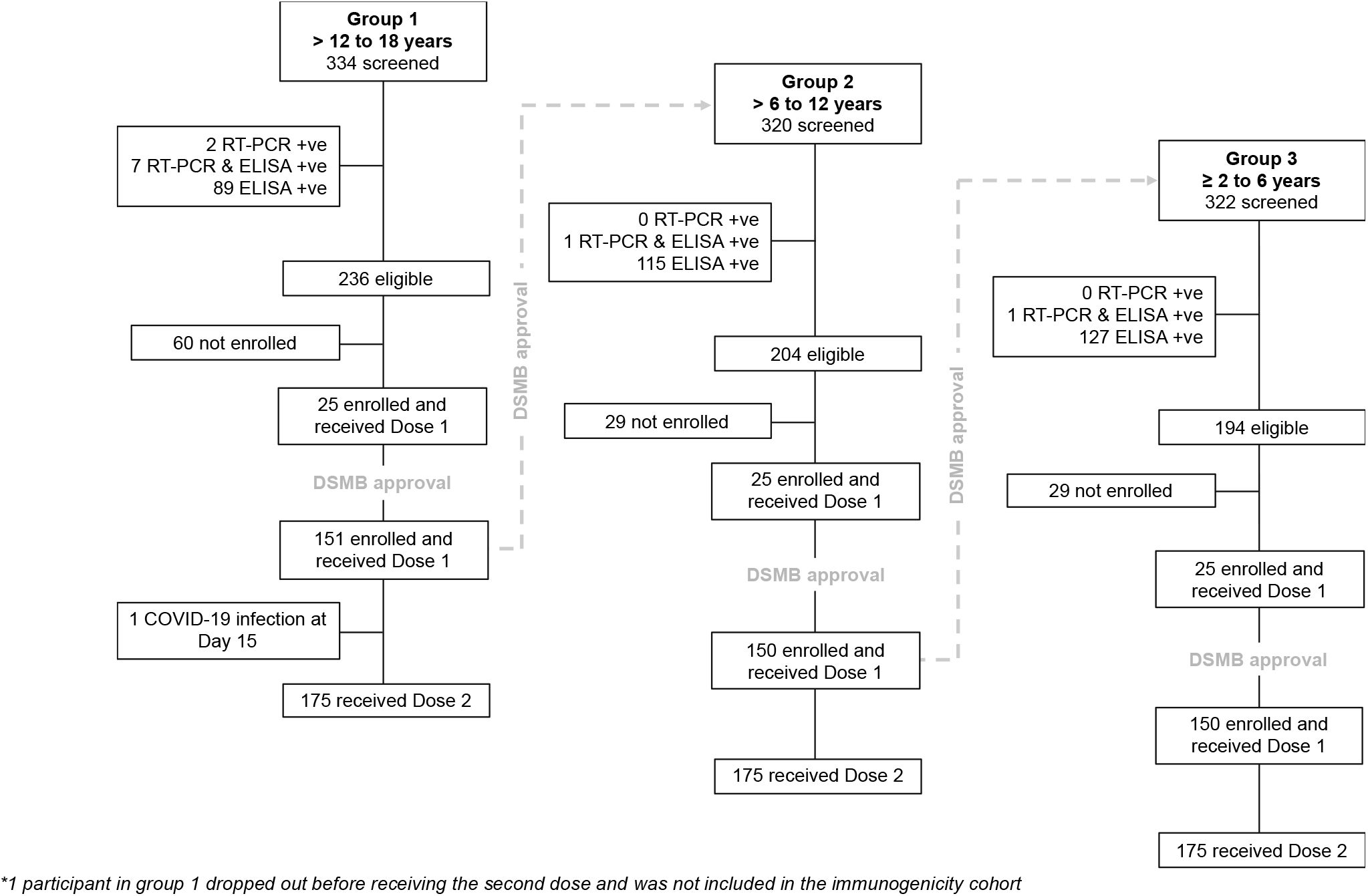
Study flow chart showing sequential enrolment of the three age groups following approval by the Data Safety Monitoring Board (DSMB).

### Safety and reactogenicity

There were no SAEs, deaths, or withdrawals due to an AE during the study except for the one case of COVID-19 infection in Group 1. There was one report of an immediate adverse event; a participant in Group 2 had mild local itching/pruritis within 2 hours of vaccination, which the investigator considered to be unlikely to be due to the vaccine. As illustrated in **Figure 2**, vaccination with BBV152 was generally well tolerated, with no statistical difference in reactogenicity profiles between the different age groups. Local reactions mainly consisted of mild injection site pain, reported by fewer than 35% of any group after the first dose, and fewer than 25% after the second dose; there were no cases of severe pain. There were few reports of other local reactions, with no consistent trend across the groups. Systemic adverse events were less frequent, especially after the second dose. After Dose 1 the most frequent systemic AE was mild-to-moderate fever, reported in 8 of 176 (5%) of Group 1 participants, 17 of 175 (10%) of Group 2, and 22 of 175 (13%) of children in Group 3. No case of severe fever was reported, and rates were all 4% or less after Dose 2.

**Figure 2.**
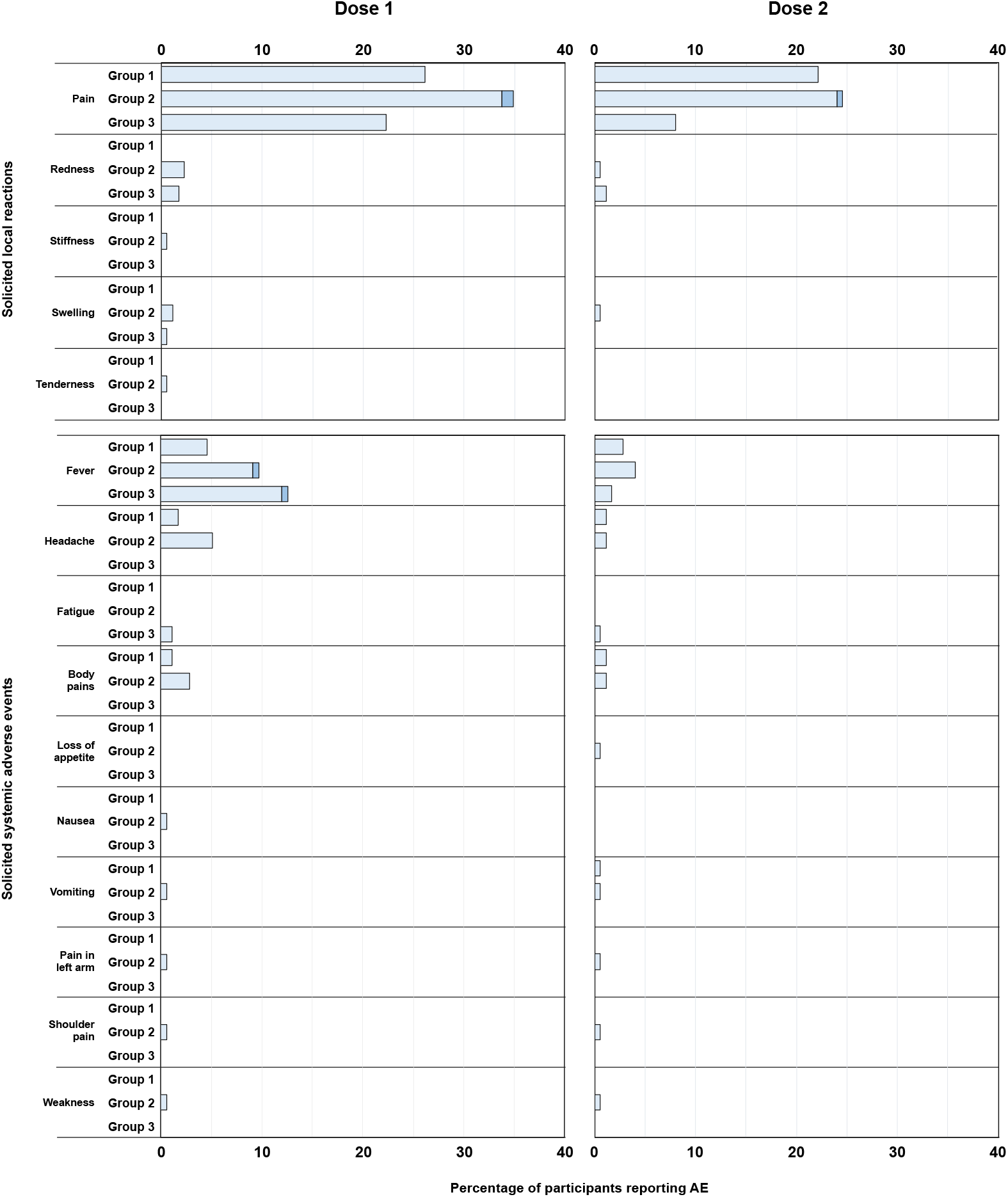
Solicited reactogenicity in the three age groups in the 7 days after the first and second doses of BBV152 in Groups 1 (> 12 – ≤ 18 year-olds), 2 (> 6 – ≤ 12 year-olds), and 3 (≥ 2 – ≤ 6 year-olds).

Unsolicited adverse events were also infrequent after vaccination, reported by 3 of 176 (2%) children in Group 1, 8 of 175 (5%) children in Group 2, and 4 of 175 (2%) children in Group 3. All unsolicited AEs, which consisted of individual reports of typical childhood complaints (*see Supplementary table 1*), were described as mild and resolved without sequelae.

### Immunogenicity

As illustrated in **Figure 3A**, there were similar immune responses measured as MNT antibody titres in all three age groups. As shown in **Figure 3A** vaccine-induced MNT responses in all groups were comparable to the GMT of 103.3 (95% CI: 50.3–202.1) from 18 BEI reference sera run in the same assay. On day 56, the GMT ratio comparing all children to adults was 0.98 (95% CI: 0.80-1.19). The response four weeks after the second dose 100% of Group 3 seroconverted, with a numerically higher GMT than either of the two older age groups, in which SCRs were 90% (158/175) and 90% (141/157) in Groups 1 and 2, respectively.

**Figure 3.**
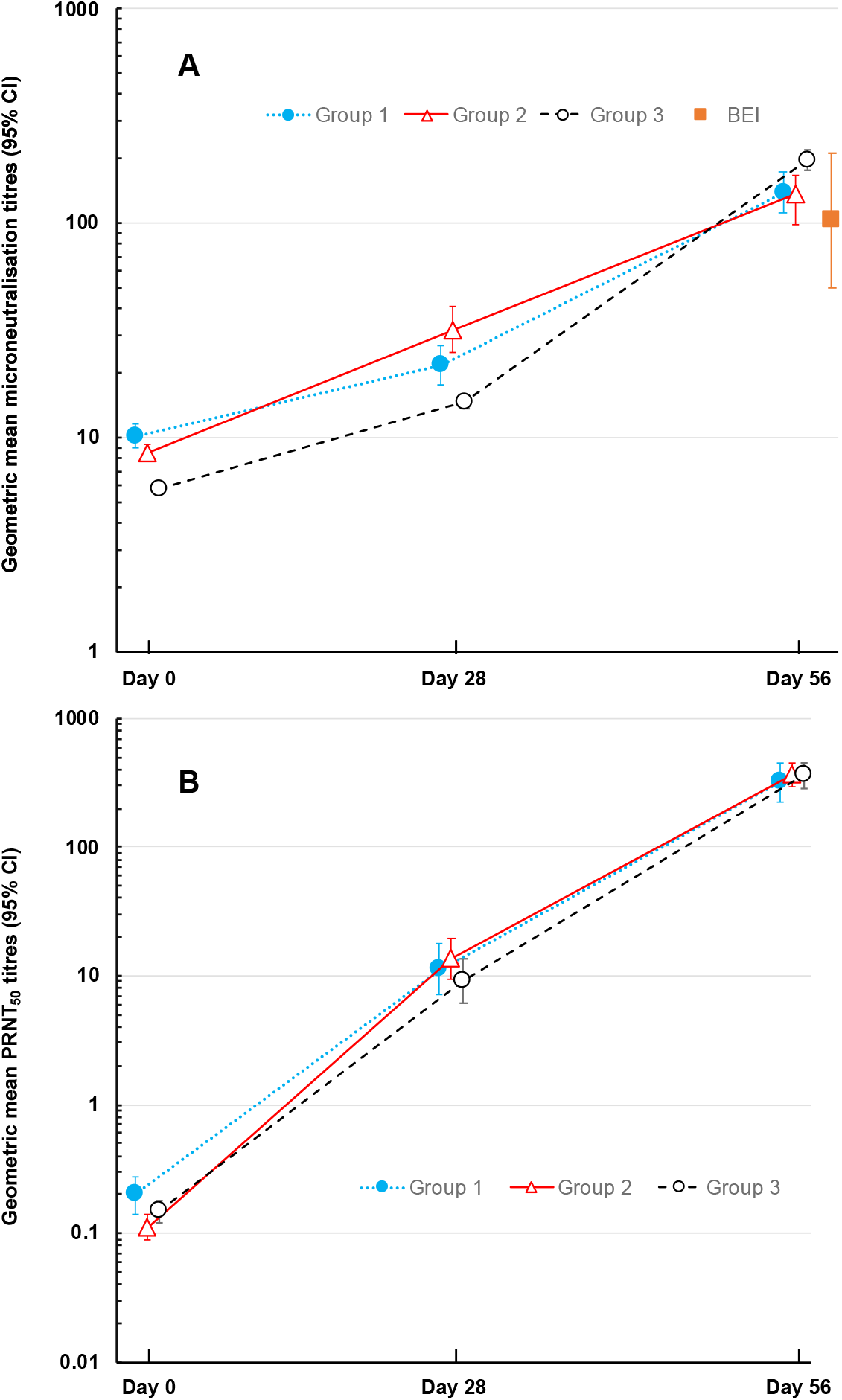
Geometric mean titres (with 95% CI) of SARS-CoV-2 neutralising antibodies measured by microneutralisation (A) or plaque reduction (B) at baseline (Day 0) and four weeks after the first (Day 28) and second (Day 56) doses of BBV152 in the three age groups: (Group 1; > 12 – ≤ 18 year-olds), (Group 2; > 6 – ≤ 12 year-olds), (Group 3; ≥ 2 – ≤ 6 year-olds). MNT_50_ GMT (95% CI) for BEI samples are shown in A.

When assessed by PRNT there was no evident difference between groups either in terms of GMTs or SCRs after two doses (**Figure 3B, Table 2**). On day 56, the GMT ratio comparing children to adults was 1.76 (95% CI: 1.32-2.33). Seroconversion after two doses achieved similarly high levels (95-98%) in all three groups four weeks after the second vaccination.

**Table 2:** Geometric mean titres (GMT) and seroconversion rates for SARS-CoV-2 neutralising antibodies measured by microneutralisation test (MNT_50_) or plaque reduction neutralisation test (PRNT_50_) – and values obtained in a phase 2 study in adults [11].

| Microneutralisation Test (MNT<sub>50</sub>) |  |  |  |  |  |  |
| --- | --- | --- | --- | --- | --- | --- |
| Day |  | Group 1 | Group 2 | Group 3 | Adults [11] | GMT Ratio (All children Vs. Adults) |
| 0 | N | 175 | 175 | 175 | - |  |
|  | GMT | 10.1 | 8.5 | 5.8 | - |  |
|  | (95% CI) | (8.9–11.5) | (7.8–9.3) | (5.7–5.9) | - |  |
| 56 | N | 175 | 157 | 173 | 177 |  |
|  | GMT | 138.8 | 137.4 | 197.6 | 160.1 | 0.98<br>(0.80 – 1.19) |
|  | (95% CI) | (111.0–173.6) | (99.1–167.5) | (176.4–221.4) | (135.8–188.8) |  |
|  | Seroconverted*, n | 158 | 141 | 173 | 171 |  |
|  | %<br>(95% CI) | **90.3**<br>(84.9–94.2) | **89.8**<br>(84.0–94.1) | **100**<br>(97.9–100) | **96.6**<br>(92.8–98.7) |  |
|  | p value vs Adult | 0.027 | 0.022 | 0.032 | - |  |
| Plaque Reduction Neutralisation Test (PRNT<sub>50</sub>) |  |  |  |  |  |  |
| 0 | N | 175 | 175 | 175 | - |  |
|  | GMT | 0.20 | 0.11 | 0.15 | - | GMT Ratio (All children Vs. Adults) |
|  | (95% CI) | (0.14–0.27) | (0.09–0.14) | (0.12–0.18) | - |  |
| 56 | N | 175 | 168 | 172 | 177 |  |
|  | GMT | 317.4 | 366.9 | 358.6 | 197.0 | 1.76<br>(1.32 – 2.33) |
|  | (95% CI) | (224.4–449.2) | (297.0–453.3) | (287.2–447.8) | (155.6–249.4) |  |
|  | Seroconverted*, n | 166 | 165 | 169 | 174 |  |
|  | %<br>(95% CI) | **94.9**<br>(90.5–97.6) | **98.2**<br>(94.9–99.6) | **98.3**<br>(95.0–99.6) | **98.3**<br>(95.1–99.6) |  |
|  | p value vs Adult | 0.13 | 1.00 | 1.00 | - |  |

\* Seroconversion defined a four-fold increase in titre over baseline at Day 0. No statistical differences in immune responses were observed between the paediatric trial participants and adults from the phase 2 clinical trial. Based on PRNT responses, neutralizing antibody responses in children were found to be superior to adults.

SCRs for neutralising antibodies after two doses as measured by PRNT were comparable in all three groups of children and the phase 2 adults, while GMTs were higher in all three groups of children (317.4, 366.9, and 358.6, respectively) than those previously reported in adults (197.0 [95% CI:155.6–249.4) (**Table 2**).

Binding IgG antibody responses against the three specific SARS-CoV-2 protein components, S-protein, RBD, and N-protein, are illustrated in **Figure 4**. These show that each of the three age groups responded in a similar manner and with comparable magnitude against the three proteins, with the exception of a lower GMT at Day 56 for N-protein in Group 3. The isotyping ratios (IgG1/IgG4) at Day 56 were substantially above 1 for all vaccinated groups, indicative of a Th1 bias (*Supplementary table 5*).

**Figure 4.**
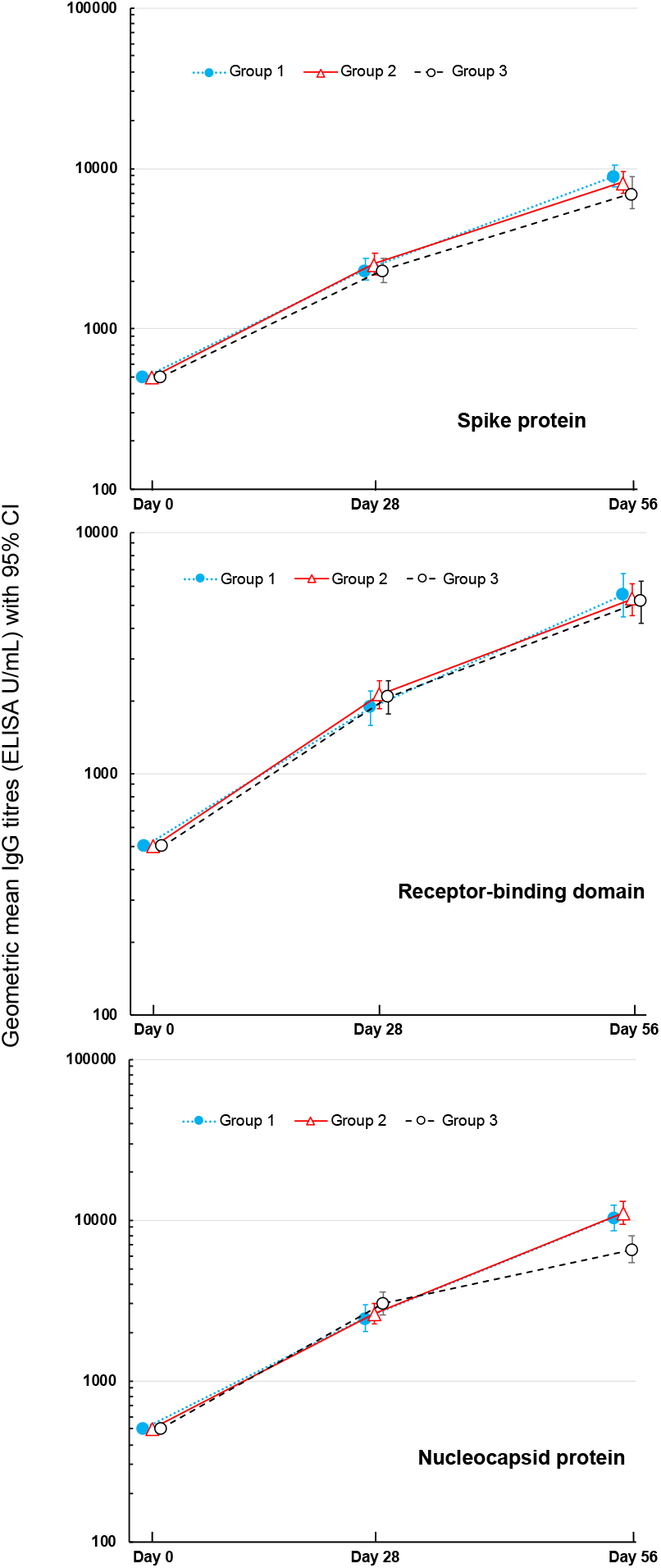
Geometric mean titres (with 95% CI) of IgG antibodies measured by ELISA against SARS-CoV-2 S-protein, receptor-binding domain (RBD) or nucleocapsid protein at baseline (Day 0) and four weeks after the first (Day 28) and second (Day 56) doses of BBV152 in the three age groups: (Group 1; > 12 – ≤ 18 year-olds), (Group 2; > 6 – ≤ 12 year-olds), (Group 3; ≥ 2 – ≤ 6 year-olds).

## DISCUSSION

We previously demonstrated that two doses of BBV152 (COVAXIN^®^) are effective in preventing COVID-19 due to SARS-CoV-2 infection in adults [3,4], and now show that the vaccine is well tolerated and immunogenic in children from 18 years down to 2 years of age. Immunogenicity analysis shows that neutralising antibody responses, measured by MNT, following two doses in a cohort of children aged from 2 to 18 years are non-inferior to those observed in adults. The PRNT GMT ratio (GMT in a paediatric subgroup / GMT in adults) had a lower limit ≥ 1, indicating superior antibodies in children when compared to adults, suggesting that BBV152 will also be efficacious in this age group. As it has been suggested that a Th1-dominant response is preferable for Covid-19 vaccines [15,16] we also demonstrated that vaccine responses were skewed towards a Th1 response with IgG1/IgG4 ratios above 1.

The study is ongoing to assess the persistence of the response and to provide a longer-term safety assessment during a period of high ongoing SARS-CoV-2 circulation. Substantial SARS-CoV-2 infection was observed in the general population (outside the trial), which may indicate post-vaccination titres from the vaccinated recipients could be slightly inflated due to natural exposure to SARS-CoV-2. The pattern of baseline neutralising antibody GMTs suggests an age-dependent increase (*Supplementary figure 1, p 7*), possibly due to older children, who presumably mix more socially, being more exposed. Only one case of symptomatic COVID-19 was detected in the study, in the oldest age-group. However, illness visits were not scheduled, and routine SARS-CoV-2 nucleic acid testing was not conducted, so asymptomatic or mild symptomatic cases of COVID-19 may have been missed.

The necessity to vaccinate children is currently unclear, but there are moves to advance in that area [10,11]. Despite high rates of COVID-19 infections in children [17], low rates of severe consequences in children suggest that this is not a priority except for children at high risk [18], i.e. those with underlying co-morbidities. However, as the pandemic is continuously evolving with emergence of new Variants of Concern [19], the future is unclear. Wise counsel would suggest that the availability of safe and effective vaccines suitable for use in children is an obvious precautionary measure [11]. An issue with all current COVID-19 vaccines is that we are unsure how effective they will be against newly emerging variants. This is particularly concerning for those vaccines tailored specifically against the S-protein of the prototype (Wuhan) virus, which is no longer the main circulating strain [20], as new variants such as the Omicron (B.1.1.529) variant continue to emerge and disperse globally [21]. Specific focus of a vaccine on one epitope or region of the virus means the immune response may be by-passed by a new strain in which the primary epitopes found on the S-protein have substantially changed. Inactivated Covid-19 vaccines have been shown to be well tolerated and immunogenic in phase 1/2 trials in Chinese children from 3 to 17 years of age [22,23]. An advantage of a whole-virion vaccine such as BBV152 is that multiple epitopes are present, as illustrated by the marked responses against S-protein, RBD, and N-protein in this study.

It has been suggested that an imbalanced Th1/Th2 immune response is a component in developing severe respiratory symptoms in some COVID-19 patients [24,25] and that vaccines against Covid-19 should develop a predominantly Th1 response [15]. As BBV152 is adjuvanted with Algel-IMDG, we assessed the IgG1/IgG4 ratio as a marker of the Th1/Th2 balance in the immune response to BBV152 [12,13].

The limitations of this study are mainly due to the ethical considerations of doing such a study in children, for which reason it was done open-label with no placebo arm. As such, we cannot compare the reactogenicity and immunogenicity results with a control arm. However, the low rates of reported reactogenicity except for injection site pain suggest that a control arm would not have revealed any medically significant issues. Transient mild-to-moderate injection site pain is the main adverse event reported by adults not only with BBV152 [3,13] but also with other COVID-19 vaccines [23,24]. Also, we have assessed only the immunogenicity of BBV152, not clinical efficacy, in children.

In conclusion, the inactivated SARS-CoV-2 vaccine, BBV152, was well tolerated and immunogenic in children from 2 to 18 years of age, with neutralising antibody responses comparable to those observed in adults in whom the vaccine has been proven to be efficacious against symptomatic and asymptomatic COVID-19.

## Data Availability

All data produced in the present study are available upon reasonable request to the authors.

## ACKNOWLEDGEMENTS

We would like to sincerely thank the volunteers, investigators, study coordinators, and healthcare workers involved in this study. We express our gratitude to the teams at Sclin Soft Technologies for their support with data management. Drs. Shashi Kanth Muni, Yuvraj Jogdand, Vinay Kumar Aileni, Jagadish Kumar, Bhargav Reddy, Ms. Sandhya Rani, Ms. Aparna Bathula, Mr. Nagaraju Pillutla, and Mr. Sunil Kumar Kantheti of Bharat Biotech participated in protocol design and clinical trial monitoring. We thank the members of the DSMB and Adjudication Committee for their continued support and guidance of this ongoing clinical program. This vaccine candidate could not have been developed without the efforts of Bharat Biotech’s Manufacturing, and Quality Control teams. We are grateful to Keith Veitch (keithveitch communications, Amsterdam, The Netherlands) for editorial assistance with the manuscript.

## Competing Interests

This work was funded by Bharat Biotech International Limited. KMV, SR, BR and SP, are employees of Bharat Biotech, with no stock options or incentives. Co-author, KE, is the Chairman and Managing Director of Bharat Biotech and owns equity in the company. RE and WB are independent clinical development and statistical consultants, respectively. NM, VNT, CS, VK, SP, NP, SRa, were principal investigators representing the study sites.

## Supplementary material

**Supplementary table 1:** Unsolicited adverse events

|  | **Group 1 (n = 176)**<br>> 12 to 18 years | **Group 2 (n = 175)**<br>> 6 to 12 years | **Group 3 (n = 175)**<br>≥ 2 to 6 years |
| --- | --- | --- | --- |
| **Any unsolicited AE** | 3 (1.7) | 8 (4.6) | 4 (2.3) |
| **Adverse event, n (%)** |  |  |  |
| Itching | - | 2 (1.1) | - |
| Abdominal pain | - | 1 (0.6) | - |
| Cold | - | - | 1 (0.6) |
| Cough | - | - | 2 (1.1) |
| Diarrhoea | 1 (0.6) | - | - |
| Eye pain | - | 1 (0.6) | - |
| Itching on hands & legs | - | 1 (0.6) | - |
| Leg pain | 1 (0.6) | - | - |
| Eye redness | 1 (0.6) | - | 1 (0.6) |
| Sneezing | - | 1 (0.6) | - |
| Stomach upset | - | 1 (0.6) | - |
| Throat pain | - | 1 (0.6) | - |

Severity of all unsolicited adverse events was described as mild

**Supplementary table 2:** Geometric mean titres (GMT) and seroconversion rates for IgG binding antibodies against SARS-CoV-2 S-protein measured by ELISA

| ELISA S-protein IgG |  |  |  |  |
| --- | --- | --- | --- | --- |
| Day |  | Group 1 | Group 2 | Group 3 |
| 0 | N | 175 | 175 | 175 |
|  | GMT<br>(95% CI) | 500<br>(500–500) | 500<br>(500–500) | 500<br>(500–500) |
| 28 | N | 175 | 175 | 175 |
|  | GMT<br>(95% CI) | 2353<br>(2018–2743) | 2557<br>(2192–2982) | 2325<br>(1963–2753) |
|  | Seroconverted*, n | 114 | 128 | 114 |
|  | %<br>(95% CI) | 65.1<br>(57.8–71.8) | 73.1<br>(66.1–79.2) | 65.1<br>(57.6–72.2) |
| 56 | N | 175 | 170 | 173 |
|  | GMT<br>(95% CI) | 8985<br>(7735–10437) | 8198<br>(7015–9580) | 7066<br>(5653–8832) |
|  | Seroconverted*, n | 170 | 162 | 142 |
|  | %<br>(95% CI) | 97.1<br>(84.9–93.9) | 95.3<br>(84.0–94.7) | 82.1<br>(75.5–87.5) |

**Supplementary table 3:** Geometric mean titres (GMT) and seroconversion rates for IgG binding antibodies against SARS-CoV-2 RBD measured by ELISA

| ELISA RBD IgG |  |  |  |  |
| --- | --- | --- | --- | --- |
| Day |  | Group 1 | Group 2 | Group 3 |
| 0 | N | 175 | 175 | 175 |
|  | GMT<br>(95% CI) | 500<br>(500–500) | 500<br>(500–500) | 500<br>(500–500) |
| 28 | N | 175 | 175 | 175 |
|  | GMT<br>(95% CI) | 1870<br>(1579–22141) | 2122<br>(1849–2436) | 2081<br>(1776–2438) |
|  | Seroconverted*, n | 98 | 112 | 110 |
|  | %<br>(95% CI) | 56.0<br>(48.6–63.2) | 64.0<br>(56.7–70.8) | 62.9<br>(55.2–70.0) |
| 56 | N | 175 | 170 | 173 |
|  | GMT<br>(95% CI) | 5513<br>(4487–6773) | 5278<br>(4529–6151) | 5149<br>(4220–6281) |
|  | Seroconverted*, n | 153 | 157 | 142 |
|  | %<br>(95% CI) | 87.4<br>(81.6–91.6) | 92.4<br>(87.3–95.6) | 82.1<br>(75.5–82.1) |

**Supplementary table 4:**
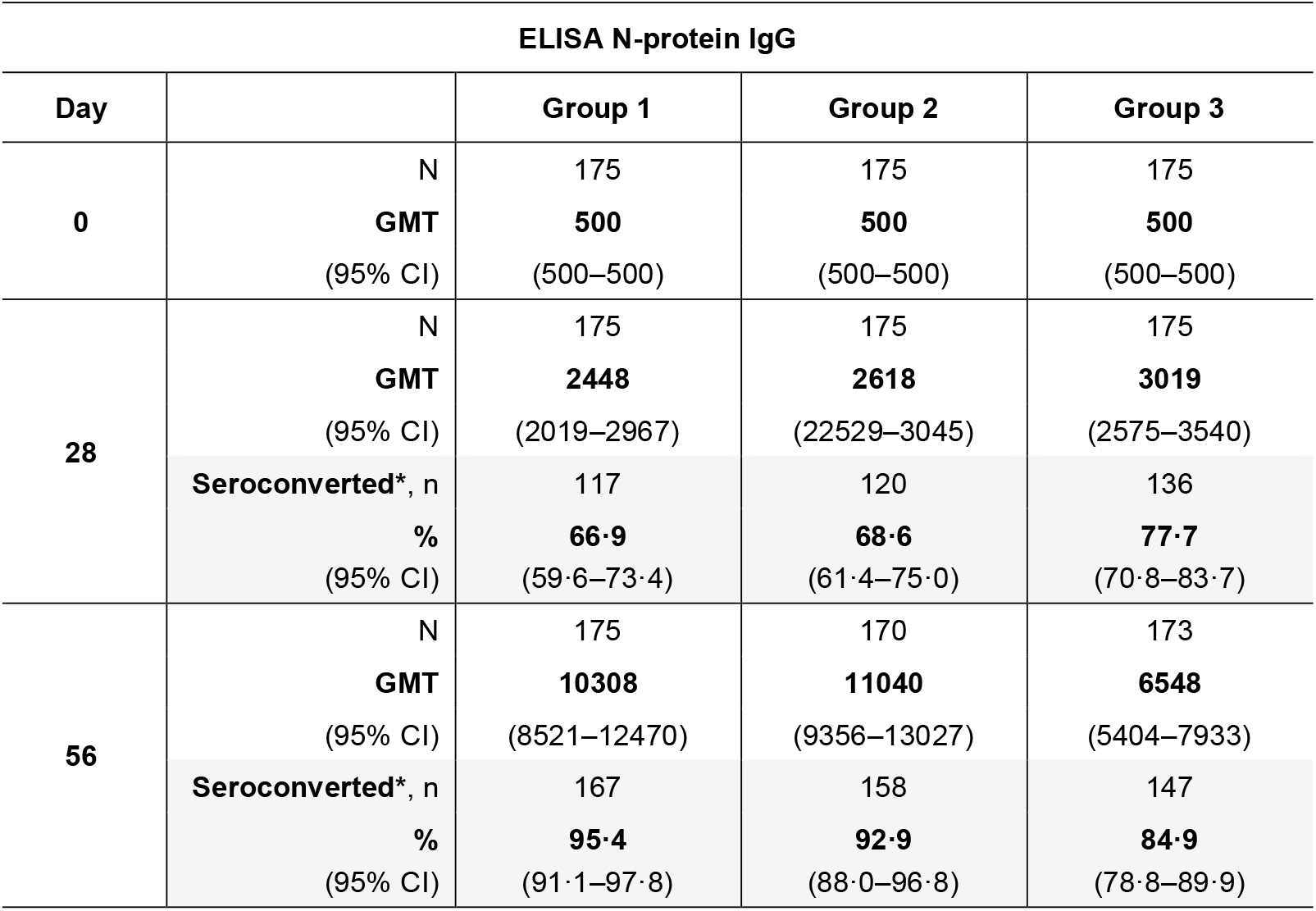
Geometric mean titres (GMT) and seroconversion rates for IgG binding antibodies against SARS-CoV-2 N-protein measured by ELISA

**Supplementary Figure 1.**
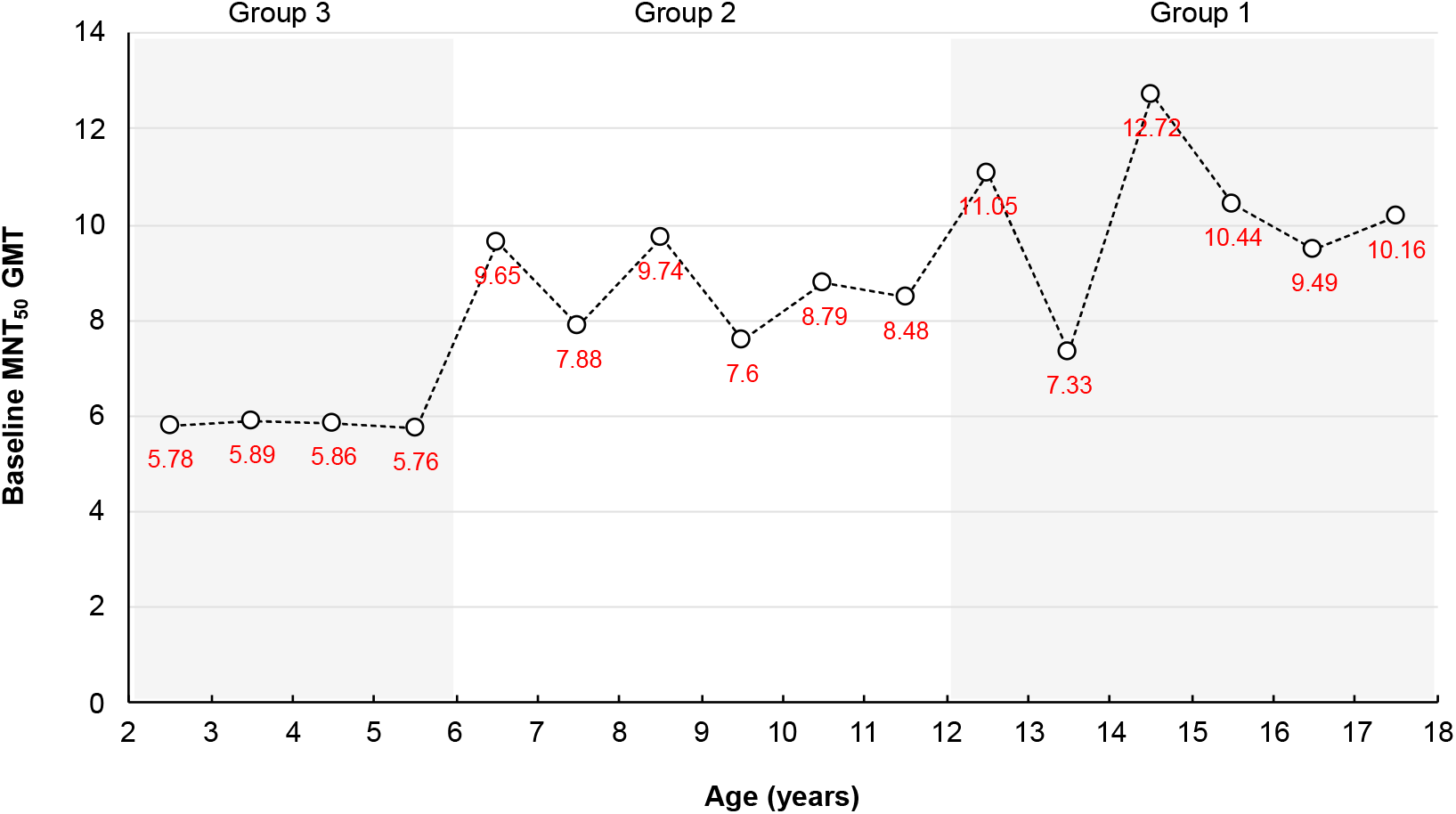
Geometric mean titres of neutralising antibodies at baseline (Day 0) measured by microneutralisation assay arranged according to mean age in one year increments

**Supplementary table 5.** Th1:Th2 index as GMT ratios of IgG1:IgG4 with 95% confidence intervals.

|  |  |  |  |
| --- | --- | --- | --- |
|  | **Th1:Th2 Index**<br>**GMTs (95% CI)** |  |  |
|  | **Group 1**<br>**13–18 years** | **Group 2**<br>**6–12 years** | **Group 3**<br>**2–6 years** |
| **Day 0** | 8.72<br>(1.38 – 55.2) | 1.54<br>(0.26 – 9.12) | 1.68<br>(0.38 – 7.37) |
| **Day 56** | 79.6<br>(304 – 1164) | 49.4<br>(21.8 – 112) | 38.1<br>(7.67 – 188) |

A Th1: Th2 ratio >1 is suggestive of vaccine induced cell-mediated immune respon

## Notes

### Clinical Trial

Registered with the Clinical Trials Registry (India) No. CTRI/2021/05/033752

### Author Declarations

1. Institutional Ethics Committee: ECR/1460/Inst/TG/2020 of Pranaam Hospital gave ethical approval for this work 2. Institutional Ethics Committee: ECR/1387/Inst/BR/2020 of All India Institute of Medical Sciences gave ethical approval for this work 3. Institutional Ethics Committee: ECR/134/Inst/KA/2013/RR-19 of Cheluvambha Hospital gave ethical approval for this work 4. Institutional Ethics Committee: ECR/608/Inst/MH/2014/RR-20 of Meditrina Institute of Medical Sciences gave ethical approval for this work. 5. Institutional Ethics Committee: ECR/1017/Inst/UP/2017/RR-21 of Prakhar Hospital gave ethical approval for this work. 6. Institutional Ethics Committee: ECR/538/Inst/DL/2014/RR-20 of All India Institute of Medical Sciences gave ethical approval for this work.

## REFERENCES

1. Crommelin DJA, Anchordoquy TJ, Volkin DB, Jiskoot W, Mastrobattista E. Addressing the cold reality of mRNA vaccine stability. J Pharm Sci 2021; 110:997–1001.

2. Ella R, Vadrevu KM, Jogdand H, et al. Safety and immunogenicity of an inactivated SARS-CoV-2 vaccine, BBV152: a double-blind, randomised, phase 1 trial. Lancet Infect Dis 2021; 21: 637–46.

3. Ella R, Reddy S, Blackwelder W, et al. Efficacy, safety, and lot-to-lot immunogenicity of an inactivated SARS-CoV-2 vaccine (BBV152): interim results of a randomised, double-blind, controlled, phase 3 trial. Lancet Published Online November 11, 2021 https://doi.org/10.1016/S0140-6736(21)02000-6

4. Desai D, Khan AR, Soneja M, et al. Effectiveness of an inactivated virus-based SARS-CoV-2 vaccine, BBV152, in India: a test-negative, case-control study. Lancet Infect Dis Published Online November 23, 2021 https://doi.org/10.1016/S1473-3099(21)00674-5

5. Ludvigsson JF. Systematic review of COVID-19 in children shows milder cases and a better prognosis than adults. Acta Paediatr 2020;109:1088–95.

6. Ladhani SN, Amin-Chowdhury Z, Davies HG, et al. COVID-19 in children: analysis of the first pandemic peak in England. Arch Dis Child 2020;105:1180–5.

7. Bi Q, Wu Y, Mei S, et al. Epidemiology and transmission of COVID-19 in 391 cases and 1286 of their close contacts in Shenzhen, China: a retrospective cohort study. Lancet Infect Dis 2020; 20: 911–9.

8. Otto SP, Day T, Arino J, et al. The origins and potential future of SARS-CoV-2 variants of concern in the evolving COVID-19 pandemic. Curr Biol 2021; 31:R918–29.

9. Mallapaty S. Will COVID become a disease of the young? Nature 2021;595: 343–4.

10. Wong BLH, Ramsay ME, Ladhani SN. Should children be vaccinated against COVID-19 now? Arch Dis Child 2021; 106: 1147–8.

11. Klass P, Ratner AJ. Vaccinating children against Covid-19 — the lessons of measles. New Engl J Med 2021; 384:589–91.

12. Kawasaki Y, Suzuki J, Sakai N, et al. Evaluation of T helper-1/-2 balance on the basis of IgG subclasses and serum cytokines in children with glomerulonephritis. Am J Kidney Dis 2004; 44:42–9.

13. Hjelholt A, Christiansen G, Sørensen US, Birkelund S. IgG subclass profiles in normal human sera of antibodies specific to five kinds of microbial antigen. Pathogens and Disease 2013; 67: 206–13.

14. Ella R, Reddy S, Jogdand H, et al. Safety and immunogenicity of an inactivated SARS-CoV-2 vaccine, BBV152: interim results from a double-blind, randomised, multicentre, phase 2 trial, and 3-month follow-up of a double-blind, randomised phase 1 trial. Lancet Infect Dis 2021; 21:950–61.

15. Jeyanathan M, SAfkhami S, Smaill F, Miller MS, Lichty BD, Xing Z. Immunological considerations for COVID-19 vaccine strategies. Nat Rev Immunol 2020;20:615–32.

16. Ewer KJ, Barrett JR, Belij-Rammerstorfer S, et al. T cell and antibody responses induced by a single dose of ChAdOx1 nCoV-19 (AZD1222) vaccine in a phase 1/2 clinical trial. Nat Med 2021; 27:270–8.

17. American Academy of Pediatrics. Children and COVID-19: State-level data report. Available at: https://www.aap.org/en/pages/2019-novel-coronavirus-covid-19-infections/children-and-covid-19-state-level-data-report/ Accessed on December 6, 2021.

18. Coronavirus Disease 2019 in Children — United States, February 12–April 2, 2020. MMWR Morb Mortal Wkl Rep 2020; 69:422–6.

19. Murano K, Guo Y, Siomi H. The emergence of SARS-CoV-2 variants threatens to decrease the efficacy of neutralizing antibodies and vaccines. Biochem Soc Trans 2021;BST20210859 Online ahead of print.

20. Nextstrain. Genomic epidemiology of SARS-CoV-2 with global subsampling. Available at: https://nextstrain.org/ncov/open/global. Accessed December 6, 2021.

21. WHO. Classification of Omicron (B.1.1.529): SARS-CoV-2 Variant of Concern. Available at: https://www.who.int/news/item/26-11-2021-classification-of-omicron-(b.1.1.529)-sars-cov-2-variant-of-concern Accessed on December 6, 2021.

22. Han B, Song Y, Li C, et al. Safety, tolerability, and immunogenicity of an inactivated SARS-CoV-2 vaccine (CoronaVac) in healthy children and adolescents: a double-blind, randomised, controlled, phase 1/2 clinical trial. Lancet Infect Dis 2021; 21:1645–53.

23. Xia SL, Zhang YT, Wang YX, et al. Safety and immunogenicity of an inactivated COVID-19 vaccine, BBIBP-CorV, in people younger than 18 years: a randomised, double-blind, controlled, phase 1/2 trial. Lancet Infect Dis 2021 Published online September 15, 2021. https://doi.org/10.1016/S1473-3099(21)00462-X

24. Pavel AB, Glickman JW, Michels JR, Kim-Schultze S, Miller RL, Guttman-Yassky, E. Th2/Th1 cytokine imbalance is associated with higher COVID-19 risk mortality. Front Genet 2021; 12:706902.

25. Gil-Etayo FJ, Suàrez-Fernández P, Cabrera-Marante O, et al. T-helper cell subset response is a determining factor in COVID-19 progression. Front Cell Infect Microbiol. 2021; 11: 624483.

26. Chapin-Bardales J, Gee J, Myers T. Reactogenicity following receipt of mRNA-based COVID-19 vaccines. JAMA 2021;325:2201–2.

27. MacDonald I, Murray SM, Reynolds CJ, Altmann DM, Boynton RJ. Comparative systematic review and meta-analysis of reactogenicity, immunogenicity and efficacy of vaccines against SARS-CoV-2. npj Vaccines 2021; 6:74

